# Head, facial and neck cooling as per-cooling modalities to improve exercise performance in the heat: A narrative review and practical applications

**DOI:** 10.1101/2021.05.31.21258125

**Authors:** Yinhang Cao, Tze-Huan Lei, Faming Wang, Bin Yang

## Abstract

It has been well established that athletic performance is greatly affected by environmental heat. Numerous studies have attempted to find reliable cooling strategies to improve athletic performance while exercising in the heat. Whole-body pre-cooling has been found to enhance endurance performance in both dry and humid heat. Nevertheless, positive physiological alternations induced by pre-cooling gradually disappear during exercise. Hence, there is a great need to find effective per-cooling strategies to improve athletic performance in the heat. Unfortunately, it’s impractical to adopt pre-cooling approaches as a per-cooling modality to improve athletic performance due to inherent issues of practicality. Thus, a narrative review was conducted to examine the impact of head, neck and facial cooling on athletic performance in the heat. Based on current evidences, head, neck and facial cooling could greatly decrease local skin temperature at those areas where cooling was applied and thereby, local perceptual sensations were greatly enhanced. Neck cooling during exercise is found effective to improve athletic performance for both endurance and team sports athletes in the heat. Besides, neck cooling is preferred over the head, facial & combined head/facial & neck cooling for both endurance and team sport athletes in the heat from a practical application viewpoint. Research is lacking on the systematically selection of per-cooling modalities to improve athletic performance based on environmental conditions and nature of the sports activity. In addition, powerful but portable head, neck and facial cooling systems are urgently required to help athletes improving performance in the heat.

**Key points:**

- Neck cooling during exercise is effective to enhance endurance performance for endurance athletes in the heat.
- Neck cooling during exercise could improve repeated sprint performance for team sport athletes in the heat.
- Head, neck & facial cooling could largely reduce local skin temperature and thereby improving local perceptual responses.
- Neck cooling is preferred over the head, facial & combined head/facial & neck cooling for both endurance and team sport athletes.
- It’s challenging to adopt facial or head cooling to cool athletes during exercise due to inherent issues of practicality.

## 1 Introduction

Current consensus [1, 2] clearly indicates that performing endurance exercise in an uncompensable heat stress environment imposes a greater thermoregulatory strain compared to cool or thermal neutral environment. This increase of thermoregulatory strain is mainly due to the inability to dissipate the metabolic heat via both dry and wet heat transfer, which consequently results in a higher core temperature (T_core_) [2]. Higher T_core_ elevates both thermal perceptual and cardiovascular strains, which subsequently causes the voluntary reduction of power output [3] or pre-mature fatigue as assessed by the time to exhaustion approach [4].

Whole-body precooling by using water-immersion technique has been found to enhance endurance performance in both dry and humid heat as assessed by time to exhaustion [5, 6] or by using the self-selected pace [7, 8]. This increase of endurance performance is mainly due to physiological alternation (i.e., lower resting core and skin temperatures) induced by precooling [5] which thereby reduces cardiovascular and perceptual strains when performing prolonged endurance exercise in the heat. These improvements in thermoregulatory function along with a lower perceptual strain greatly enhance endurance performance in an uncompensable heat stress environment [9]. Nevertheless, the major limitations associated with whole-body pre-cooling are: time consuming and lack of real world-application because this requires a prolonged time to achieve the desired results. Also, the majority of reported whole-body cooling strategies are difficult to be adopted during exercise because such strategies are mainly designed for cooling inactive athletes. Furthermore, it’s common that the desired physiological status (e.g., enhanced heat storage capacity) induced by the use of a specific precooling strategy disappears rapidly during exercise and thereby almost no superior benefits could be registered on the alleviation of athletes’ thermal strain as compared to control trials. Recently the use of per-cooling strategy to cool athletes during exercise became of greater interest. In fact, when performing exercise in an uncompensable heat stress environment (i.e., humid heat), per cooling is just as important as pre-cooling as it is effective to reduce the rise of T_core_ during exercise [10, 11] and thereby reducing the risk of heat related injuries for all athletes.

To date, per-cooling has been extensively used for reducing athletes’ thermal strain during exercise because there is a great need to help athletes dissipate metabolic heat during the exercise compared to resting or warming-up conditions. Per-cooling strategies have often been applied to local body regions such as the torso, head, face, and the neck. Cooling the head and neck regions are effective to mitigate perceptual strain and enhance physical performance in the heat [12–16]. Those two cooling interventions are based on the facts that the head/face and neck regions of the human body have greater alliesthesial response than the rest of the body, which can immediately result in the reduction of whole-body thermal discomfort when cold stimulus is applied to mild-heat stress individuals [17].This reduction of thermal perceptions subsequently enhances both endurance and repeated sprint ability in the heat without altering the thermoregulatory response [18]. Furthermore, the cooling areas of those regions are relatively small yet achieving similar ergogenic benefits as using whole-body pre-cooling using the water immersion approach. Due to these reasons, both cooling interventions (Head and neck cooling) are considered having more real-world implications than the traditional whole-body cooling intervention as it can provide both pre and per-cooling effect for endurance athletes as well as for team sport athletes. In addition, the cooling device for both neck and head/face cooling are easy to wear with minimal weight bearing, which can be beneficial for both team sport and endurance athletes, where the weight of the equipment may have a detrimental outcome on physical performance.

Although previous investigations [10, 13–15, 18] have eloquently elucidated that both head/face and neck cooling interventions can effectively enhance performance in the heat, those investigations do not specifically address whether those interventions can be applied to both team sport and endurance athletes. Furthermore, limited studies are available to describe the physiological and performances differences between such cooling interventions. Identifying a “one size fits all” cooling intervention for all athletes is necessary for the next upcoming sporting event such as the 2021 Tokyo Olympic games as it can enhance physical performance whilst reducing the thermoregulatory strain in an uncompensable heat stress environment. Due to the shortcoming investigation between head, facial and neck cooling on physical performance, the purpose of this narrative review is to investigate which cooling intervention is ideal for both endurance and team sport athletes in an uncompensable heat stress environment. In addition, this narrative review also comprehensively overviews the thermoregulatory and perceptual response of each cooling intervention and thus provides valuable recommendation for the general public as which cooling intervention is preferred when performing daily living activity in the summer time. Lastly, we hope by highlighting the unexplored issues would stimulate more mechanistic research in this area.

## 2 Literature search methods and considerations

Literature searching was performed within the databases PubMed, MEDLINE, Scopus, Google Scholar and ProQuest up to May 2021. Keywords used for searching included neck cooling, carotid cooling, head cooling, facial cooling, exercise, per-cooling, pre-cooling, post-cooling, personal cooling, endurance performance, repeated sprint and time trial. Studies were included if one of the below criteria met:

- Participants were described as ‘healthy’ or ‘active’ and no known diseases that could affect the exercise performance or thermoregulation.
- Studies were performed at an ambient temperature of ≥ 20 °C;
- Studies examined at least a type of cooling strategy that being applied to the head, face or the neck or any possible combination of these three body regions;
- Studies reported either physiological responses or perceptual responses or both;
- Studies that were published in the English language in peer-review journals, conference proceedings or published theses.

We did not include studies that used head, face, or/and neck cooling modalities on such occupational activities such as firefighting, farming, or construction work. Any study that did not report the test condition was also excluded. Two old studies published in 1970s and 1980s were excluded as well because we were not able to access to the full-text.

Our search found 32 references on “neck cooling” AND “exercise” AND “healthy”, 22 results on “face cooling” AND “exercise” AND “healthy”, 22 results on “head cooling” AND “exercise” AND “healthy”, 15 results on “head and face cooling” AND “exercise” AND “healthy”, 9 results on “head and neck cooling” AND “exercise” AND “healthy” and 7 results on “face and neck cooling” AND “exercise”. Of those 107 studies, 64 references did not address the areas of interest. In Google Scholar and ProQuest databases, 5 relevant conference proceeding articles/extended abstracts and 1 relevant published thesis were found. Hence, a total of 49 studies have been included in this narrative review.

Figure 1 depicts the co-occurrence map of the most frequent used keywords in the abstract and the full text of the 49 studies. The co-occurrence map illustrates the extent of keywords occurring simultaneously in the reviewed keyword volume. The connection between any two simultaneous keywords is represented by the network, while the frequency of a particular keyword is represented by its circle size. The dense network demonstrates that keywords such as “male”, “female”, “face”, “head”, “neck”, “body temperature”, “skin temperature”, and “heart rate” are used frequently and are correlated with almost every other keyword. Furthermore, most studies chose “young adult” subjects as their investigated groups.

**Figure 1.**
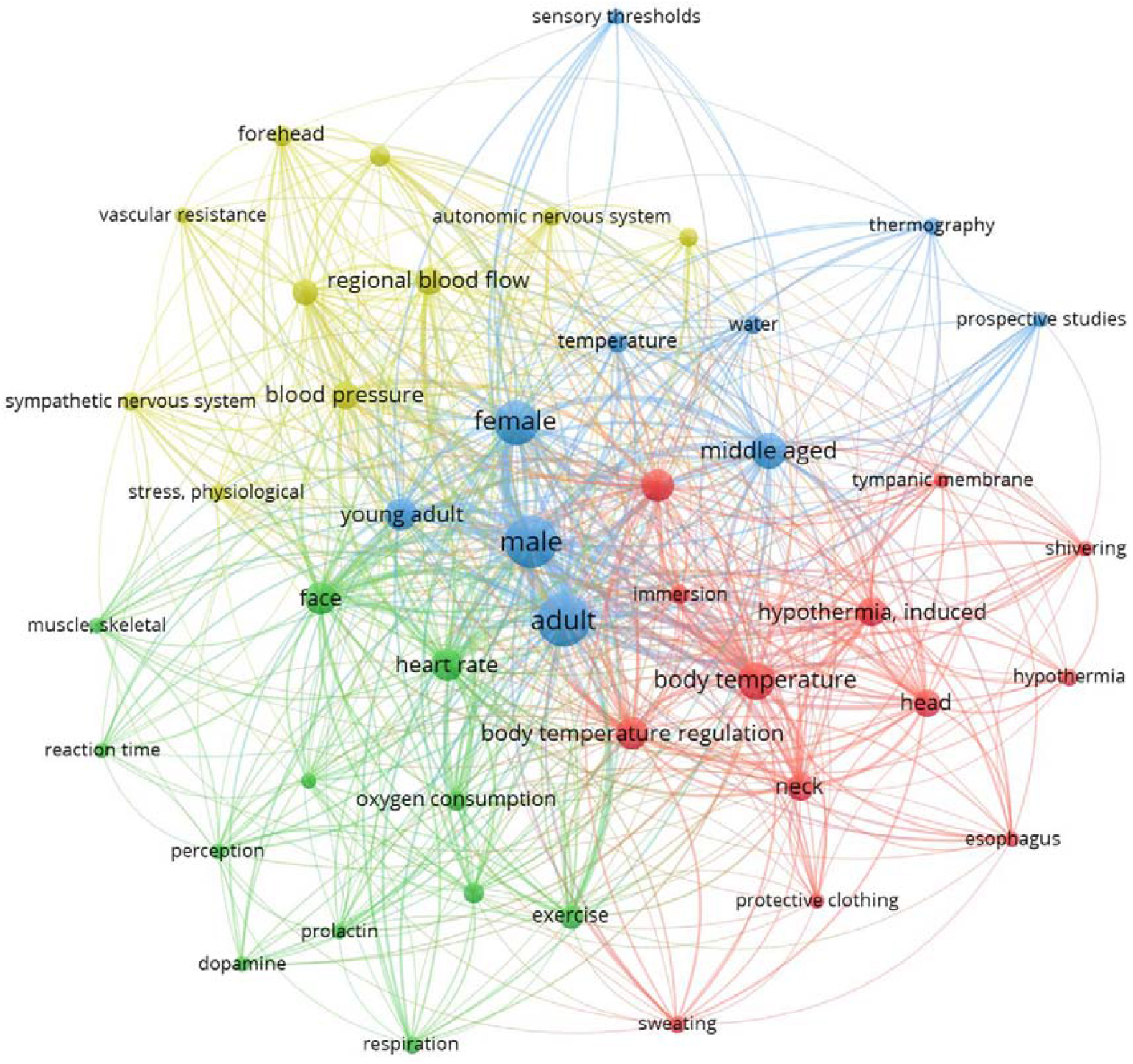
Co-occurrence map of frequently used keywords from the 49 published references.

## 3 Neck cooling and exercise performance

Table 1 shows the published studies on neck cooling and its performance on athletes while exercising in the heat [13–15, 18–36]. The most often used neck cooling approaches in the 23 documented literatures mainly include cooling collars, wet towels and ice bags. First, the use of neck cooling could greatly decrease the local skin temperature at the neck and thereby local thermal sensation at the neck was also significantly improved at the neck region, despite six out of 23 studies did not report the local neck temperature. Neck cooling via using a neck collar has been found to enhance both endurance and team sport performance in the heat [13–15, 18, 19, 28–30]. Hence, cooling the neck may be the optimal site than the face or the head. This is evidenced by the fact that time to exhaustion, self-paced performance and repeated sprint ability [13–15, 18, 19] are all greater when wearing the cold neck collar compared to no cooling trial. This increase in physical performance with neck cooling may be due to the neck region has high alliesthesial response and close proximity to the thermoregulatory center, namely the insular cortex and so any cold stimulus can directly result in the immediate change of local thermal sensation (TS) and hence the rate of perceived exertion. This therefore results in the increase of self-selected pace or extend the time to reach volitional exhaustion. The increase of endurance performance with wearing cold neck collar is mainly perceptually mediated and its thermoregulatory response is different between time to exhaustion and self-selected pace approach.

**Table 1.**
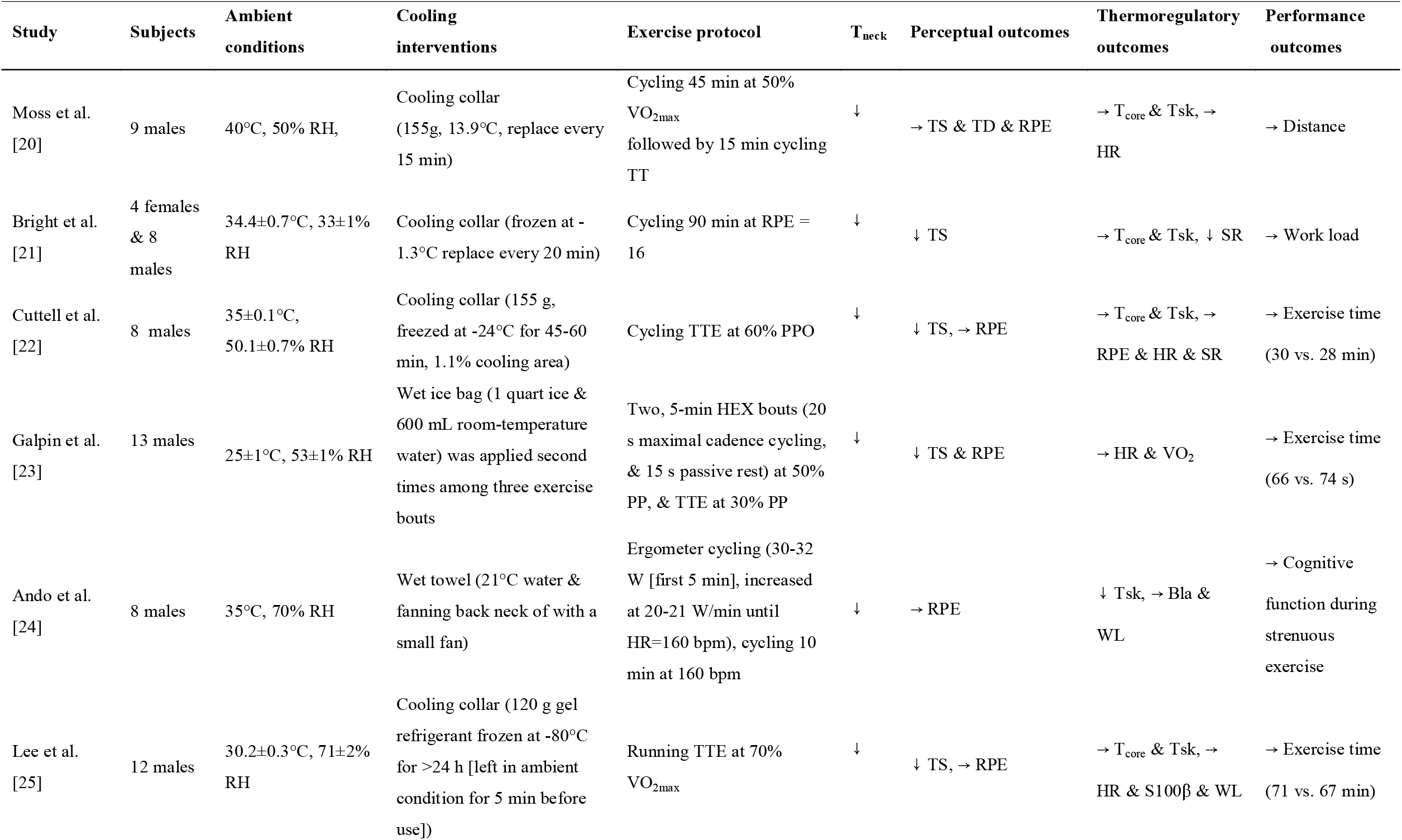
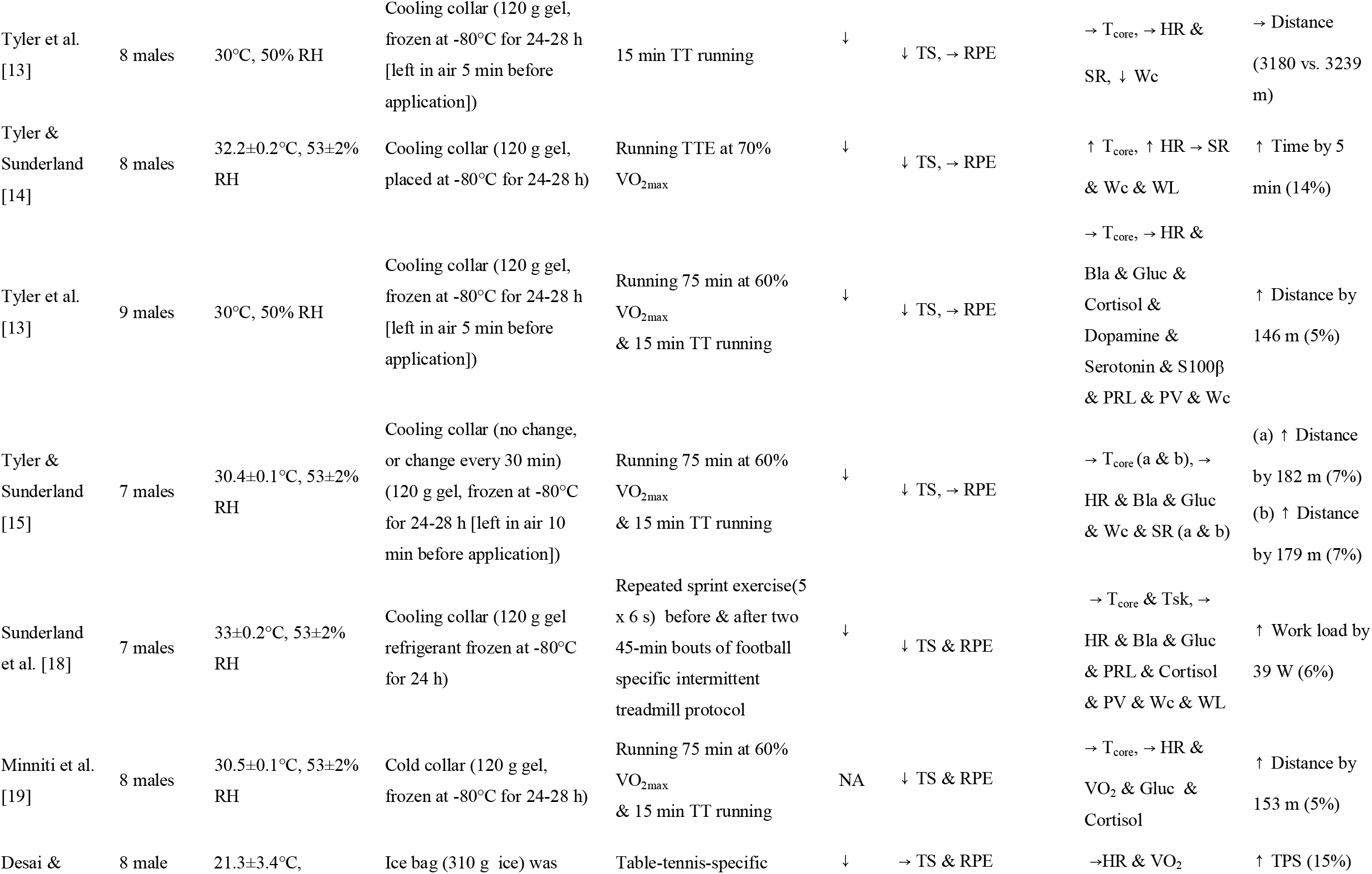
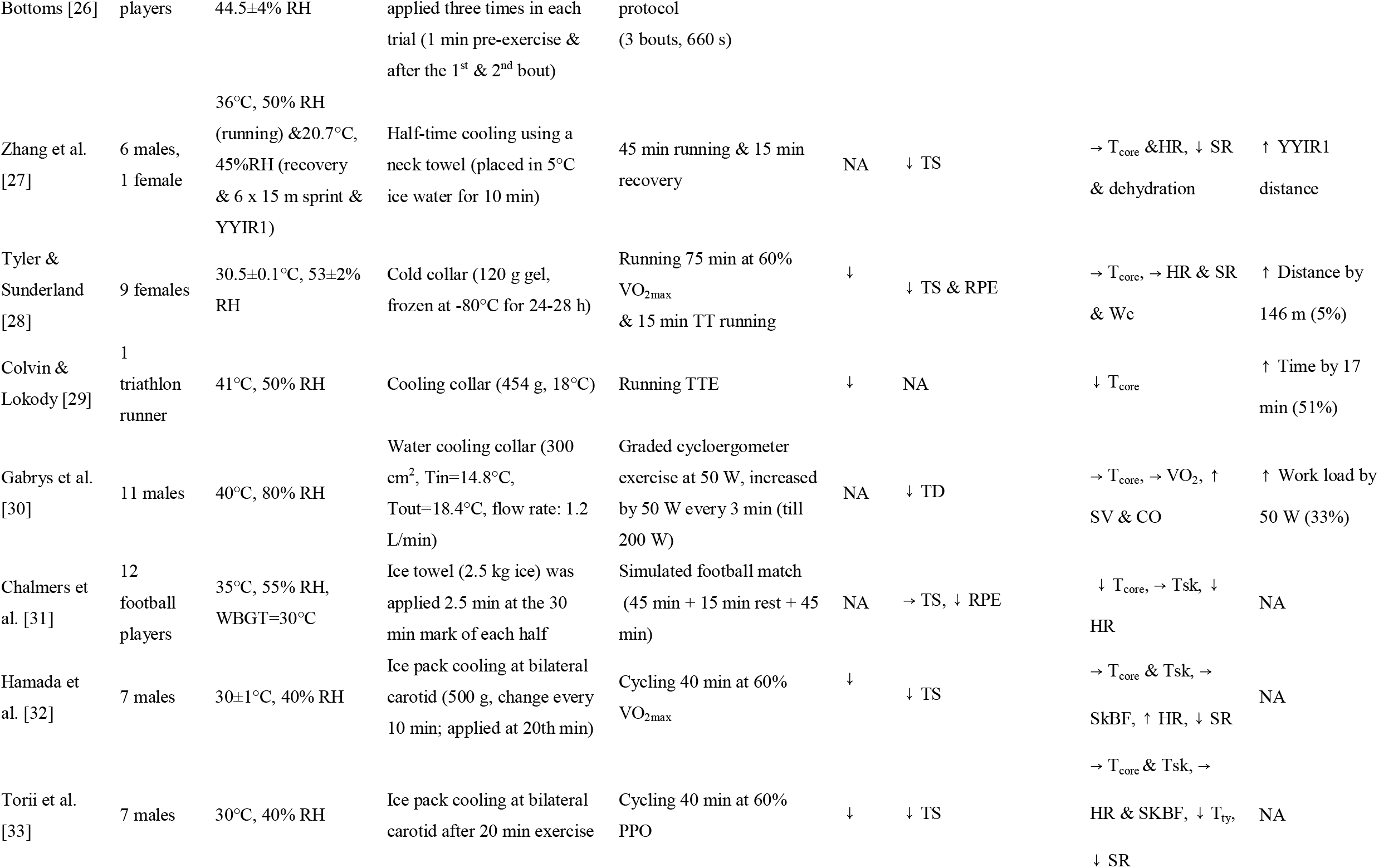
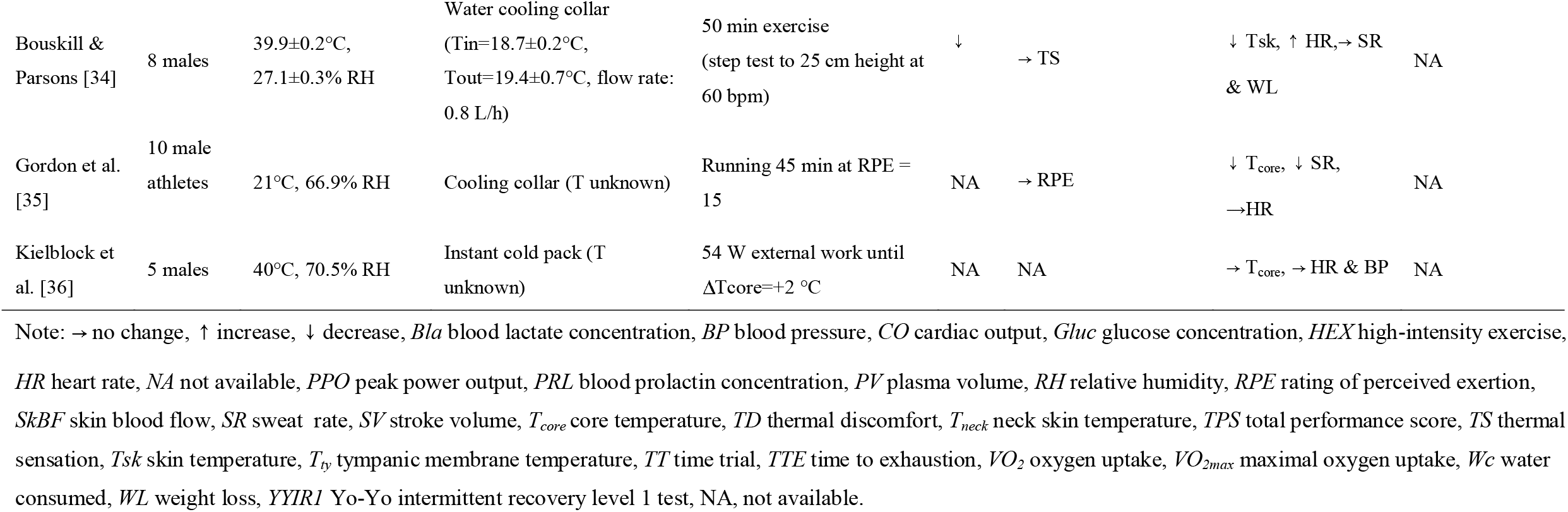
Effect of neck cooling on thermoregulatory responses and physical performance in the heat.

| Study | Subjects | Ambient conditions | Cooling interventions | Exercise protocol | T<sub>neck</sub> | Perceptual outcomes | Thermoregulatory outcomes | Performance outcomes |
| --- | --- | --- | --- | --- | --- | --- | --- | --- |
| Moss et al. [20] | 9 males | 40°C, 50% RH, | Cooling collar (155g, 13.9°C, replace every 15 min) | Cycling 45 min at 50% VO<sub>2max</sub> followed by 15 min cycling TT | ↓ | → TS & TD & RPE | → T<sub>core</sub> & Tsk, → HR | → Distance |
| Bright et al. [21] | 4 females & 8 males | 34.4±0.7°C, 33±1% RH | Cooling collar (frozen at -1.3°C replace every 20 min) | Cycling 90 min at RPE = 16 | ↓ | ↓ TS | → T<sub>core</sub> & Tsk, ↓ SR | → Work load |
| Cuttell et al. [22] | 8 males | 35±0.1°C, 50.1±0.7% RH | Cooling collar (155 g, freezed at -24°C for 45-60 min, 1.1% cooling area) | Cycling TTE at 60% PPO | ↓ | ↓ TS, → RPE | → T<sub>core</sub> & Tsk, → RPE & HR & SR | → Exercise time (30 vs. 28 min) |
| Galpin et al. [23] | 13 males | 25±1°C, 53±1% RH | Wet ice bag (1 quart ice & 600 mL room-temperature water) was applied second times among three exercise bouts | Two, 5-min HEX bouts (20 s maximal cadence cycling, & 15 s passive rest) at 50% PP, & TTE at 30% PP | ↓ | ↓ TS & RPE | → HR & VO<sub>2</sub> | → Exercise time (66 vs. 74 s) |
| Ando et al. [24] | 8 males | 35°C, 70% RH | Wet towel (21°C water & fanning back neck of with a small fan) | Ergometer cycling (30-32 W [first 5 min], increased at 20-21 W/min until HR=160 bpm), cycling 10 min at 160 bpm | ↓ | → RPE | ↓ Tsk, → Bla & WL | → Cognitive function during strenuous exercise |
| Lee et al. [25] | 12 males | 30.2±0.3°C, 71±2% RH | Cooling collar (120 g gel refrigerant frozen at -80°C for >24 h [left in ambient condition for 5 min before use]) | Running TTE at 70% VO<sub>2max</sub> | ↓ | ↓ TS, → RPE | → T<sub>core</sub> & Tsk, → HR & S100β & WL | → Exercise time (71 vs. 67 min) |
| Tyler et al. [13] | 8 males | 30°C, 50% RH | Cooling collar (120 g gel, frozen at -80°C for 24-28 h [left in air 5 min before application]) | 15 min TT running | ↓ | ↓ TS, → RPE | → T<sub>core</sub>, → HR & SR, ↓ Wc | → Distance (3180 vs. 3239 m) |
| Tyler & Sunderland [14] | 8 males | 32.2±0.2°C, 53±2% RH | Cooling collar (120 g gel, placed at -80°C for 24-28 h) | Running TTE at 70% VO<sub>2max</sub> | ↓ | ↓ TS, → RPE | ↑ T<sub>core</sub>, ↑ HR → SR & Wc & WL | ↑ Time by 5 min (14%) |
| Tyler et al. [13] | 9 males | 30°C, 50% RH | Cooling collar (120 g gel, frozen at -80°C for 24-28 h [left in air 5 min before application]) | Running 75 min at 60% VO<sub>2max</sub> & 15 min TT running | ↓ | ↓ TS, → RPE | → T<sub>core</sub>, → HR & Bla & Gluc & Cortisol & Dopamine & Serotonin & S100β & PRL & PV & Wc | ↑ Distance by 146 m (5%) |
| Tyler & Sunderland [15] | 7 males | 30.4±0.1°C, 53±2% RH | Cooling collar (no change, or change every 30 min) (120 g gel, frozen at -80°C for 24-28 h [left in air 10 min before application]) | Running 75 min at 60% VO<sub>2max</sub> & 15 min TT running | ↓ | ↓ TS, → RPE | → T<sub>core</sub> (a & b), → HR & Bla & Gluc & Wc & SR (a & b) | (a) ↑ Distance by 182 m (7%)<br>(b) ↑ Distance by 179 m (7%) |
| Sunderland et al. [18] | 7 males | 33±0.2°C, 53±2% RH | Cooling collar (120 g gel refrigerant frozen at -80°C for 24 h) | Repeated sprint exercise(5 x 6 s) before & after two 45-min bouts of football specific intermittent treadmill protocol | ↓ | ↓ TS & RPE | → T<sub>core</sub> & Tsk, → HR & Bla & Gluc & PRL & Cortisol & PV & Wc & WL | ↑ Work load by 39 W (6%) |
| Minniti et al. [19] | 8 males | 30.5±0.1°C, 53±2% RH | Cold collar (120 g gel, frozen at -80°C for 24-28 h) | Running 75 min at 60% VO<sub>2max</sub> & 15 min TT running | NA | ↓ TS & RPE | → T<sub>core</sub>, → HR & VO<sub>2</sub> & Gluc & Cortisol | ↑ Distance by 153 m (5%) |
| Desai & | 8 male | 21.3±3.4°C, | Ice bag (310 g ice) was | Table-tennis-specific | ↓ | → TS & RPE | →HR & VO<sub>2</sub> | ↑ TPS (15%) |
| Bottoms [26] | players | 44.5±4% RH | applied three times in each trial (1 min pre-exercise & after the 1<sup>st</sup> & 2<sup>nd</sup> bout) | protocol (3 bouts, 660 s) |  |  |  |  |
| Zhang et al. [27] | 6 males, 1 female | 36°C, 50% RH (running) & 20.7°C, 45%RH (recovery & 6 x 15 m sprint & YYIR1) | Half-time cooling using a neck towel (placed in 5°C ice water for 10 min) | 45 min running & 15 min recovery | NA | ↓ TS | → T<sub>core</sub> & HR, ↓ SR & dehydration | ↑ YYIR1 distance |
| Tyler & Sunderland [28] | 9 females | 30.5±0.1°C, 53±2% RH | Cold collar (120 g gel, frozen at -80°C for 24-28 h) | Running 75 min at 60% VO<sub>2max</sub> & 15 min TT running | ↓ | ↓ TS & RPE | → T<sub>core</sub>, → HR & SR & Wc | ↑ Distance by 146 m (5%) |
| Colvin & Lokody [29] | 1 triathlon runner | 41°C, 50% RH | Cooling collar (454 g, 18°C) | Running TTE | ↓ | NA | ↓ T<sub>core</sub> | ↑ Time by 17 min (51%) |
| Gabrys et al. [30] | 11 males | 40°C, 80% RH | Water cooling collar (300 cm<sup>2</sup>, Tin=14.8°C, Tout=18.4°C, flow rate: 1.2 L/min) | Graded cycloergometer exercise at 50 W, increased by 50 W every 3 min (till 200 W) | NA | ↓ TD | → T<sub>core</sub>, → VO<sub>2</sub>, ↑ SV & CO | ↑ Work load by 50 W (33%) |
| Chalmers et al. [31] | 12 football players | 35°C, 55% RH, WBGT=30°C | Ice towel (2.5 kg ice) was applied 2.5 min at the 30 min mark of each half | Simulated football match (45 min + 15 min rest + 45 min) | NA | → TS, ↓ RPE | ↓ T<sub>core</sub>, → Tsk, ↓ HR | NA |
| Hamada et al. [32] | 7 males | 30±1°C, 40% RH | Ice pack cooling at bilateral carotid (500 g, change every 10 min; applied at 20th min) | Cycling 40 min at 60% VO<sub>2max</sub> | ↓ | ↓ TS | → T<sub>core</sub> & Tsk, → SkBF, ↑ HR, ↓ SR | NA |
| Torii et al. [33] | 7 males | 30°C, 40% RH | Ice pack cooling at bilateral carotid after 20 min exercise | Cycling 40 min at 60% PPO | ↓ | ↓ TS | → T<sub>core</sub> & Tsk, → HR & SKBF, ↓ T<sub>ty</sub>, ↓ SR | NA |
| Bouskill & Parsons [34] | 8 males | 39.9±0.2°C,<br>27.1±0.3% RH | Water cooling collar<br>($T_{in}=18.7\pm0.2^{\circ}\text{C}$,<br>$T_{out}=19.4\pm0.7^{\circ}\text{C}$, flow rate:<br>0.8 L/h) | 50 min exercise<br>(step test to 25 cm height at<br>60 bpm) | ↓ | → TS | ↓ Tsk, ↑ HR, → SR<br>& WL | NA |
| Gordon et al. [35] | 10 male<br>athletes | 21°C, 66.9% RH | Cooling collar (T unknown) | Running 45 min at RPE =<br>15 | NA | → RPE | ↓ $T_{core}$, ↓ SR,<br>→ HR | NA |
| Kielblock et al. [36] | 5 males | 40°C, 70.5% RH | Instant cold pack (T<br>unknown) | 54 W external work until<br>$\Delta T_{core}=+2^{\circ}\text{C}$ | NA | NA | → $T_{core}$, → HR & BP | NA |

Note: → no change, ↑ increase, ↓ decrease, *Bla* blood lactate concentration, *BP* blood pressure, *CO* cardiac output, *Gluc* glucose concentration, *HEX* high-intensity exercise, *HR* heart rate, *NA* not available, *PPO* peak power output, *PRL* blood prolactin concentration, *PV* plasma volume, *RH* relative humidity, *RPE* rating of perceived exertion, *SkBF* skin blood flow, *SR* sweat rate, *SV* stroke volume, $T_{core}$ core temperature, *TD* thermal discomfort, $T_{neck}$ neck skin temperature, *TPS* total performance score, *TS* thermal sensation, *Tsk* skin temperature, $T_{ty}$ tympanic membrane temperature, *TT* time trial, *TTE* time to exhaustion, $VO_2$ oxygen uptake, $VO_{2max}$ maximal oxygen uptake, *Wc* water consumed, *WL* weight loss, *YYIRI* Yo-Yo intermittent recovery level 1 test, *NA*, not available.

When assessing endurance performance using the time to exhaustion approach, time to volitional exhaustion is significantly longer with the neck cooling collar compared to the control trial. However, this also corresponds to a higher core temperature at the time to exhaustion[14], leading to a conclusion that neck cooling may increase the chance of heat related illness as it masks the extent of thermal strain being perceived. However, this statement is in fact debatable as this approach deprives our thermoregulatory behavior, which has unlimited capacity to regulate our body temperature during exercise. Furthermore, time to exhaustion has poor face validity and cannot represent the actual endurance event due to its open-end design. Studies from self-selected pace indicated that neck cooling can enhance the overall power output without altering core temperature, heart rate and neuroendocrinological response [13, 15]. This indicate that using our behavior response is able to counteract the false signal induced by the neck cooling and thereby prevent the excessive rise of core temperature in the heat. Therefore, neck cooling is an effective per cooling method to enhance endurance performance in the heat. However, it is worth noting that neck cooling may not work well if the duration of exercise is less than 15 min as Tyler et al. [13] showed that neck cooling was not effective to enhance physical performance in a 15 min time trial.

It is also worth mentioning that some studies (13 out of 23, Table 1) [13, 20–25, 31–36] indicate that neck cooling per se does not render any ergogenic benefit when performing prolonged exercise in the heat. Such great discrepancy can be attributed to the intensity of the cooling is not sufficient enough to alter the rate of perceived exertion (RPE), which is commonly believed as the important perceptual marker to determine central fatigue in the heat as it directly correlated with our arousal level [68]. In particular, the α/β wave ratio was elevated during hyperthermia and this has been linked to reduce our arousal level and contributes to the inability to maintain the desired power output during prolonged exercise in the heat.

## 4 Head cooling and exercise performance

A summary of 11 published literatures on head cooling and its impact on sports and exercise performance is listed in Table 2 [16, 37–45]. Interestingly, only four studied examined the local head temperature and all found that there was a pronounced temperature reduction at the head. Over half (6 out of 11 studies) studies did not examine the perceptual responses of athletes during head cooling. It could be inferred from 4 published work [38–41] that head cooling could potentially enhance endurance performance in the heat [38–41] but whether it can be applied to both endurance events and team sport setting requires further investigation. This is given by the fact that current studies (see Table 2) have yet to investigate whether head cooling can potentially enhance repeated sprint ability for team sport athletes in any given thermal environment. Furthermore, whether head cooling is able to enhance endurance performance in the heat still remains equivocal as the majority of studies revealed that head cooling renders no ergogenic benefits in the heat. Presently, there are very limited studies available on the effect of head cooling on self-selected pace in the heat. Such conflicting findings between the previous studies are once again related to the intensity of the cooling and this is directly related to the cooling materials being used inside the cooling cap or cooling helmet. It is possible that most of the previous studies have employed low cooling intensity for their study design and thus it was insufficient to alter the RPE response during exercise in the heat. On the other hand, some studies (Table 2) have revealed that when the cooling intensity is able to attenuate the rise of RPE, the increase of physical performance ensues [16, 40]. Therefore, it is believed that head cooling with stronger cooling intensity could potentially alter our arousal level and thereby affecting our physical performance. However, this specific notion requires further justification.

**Table 2.**
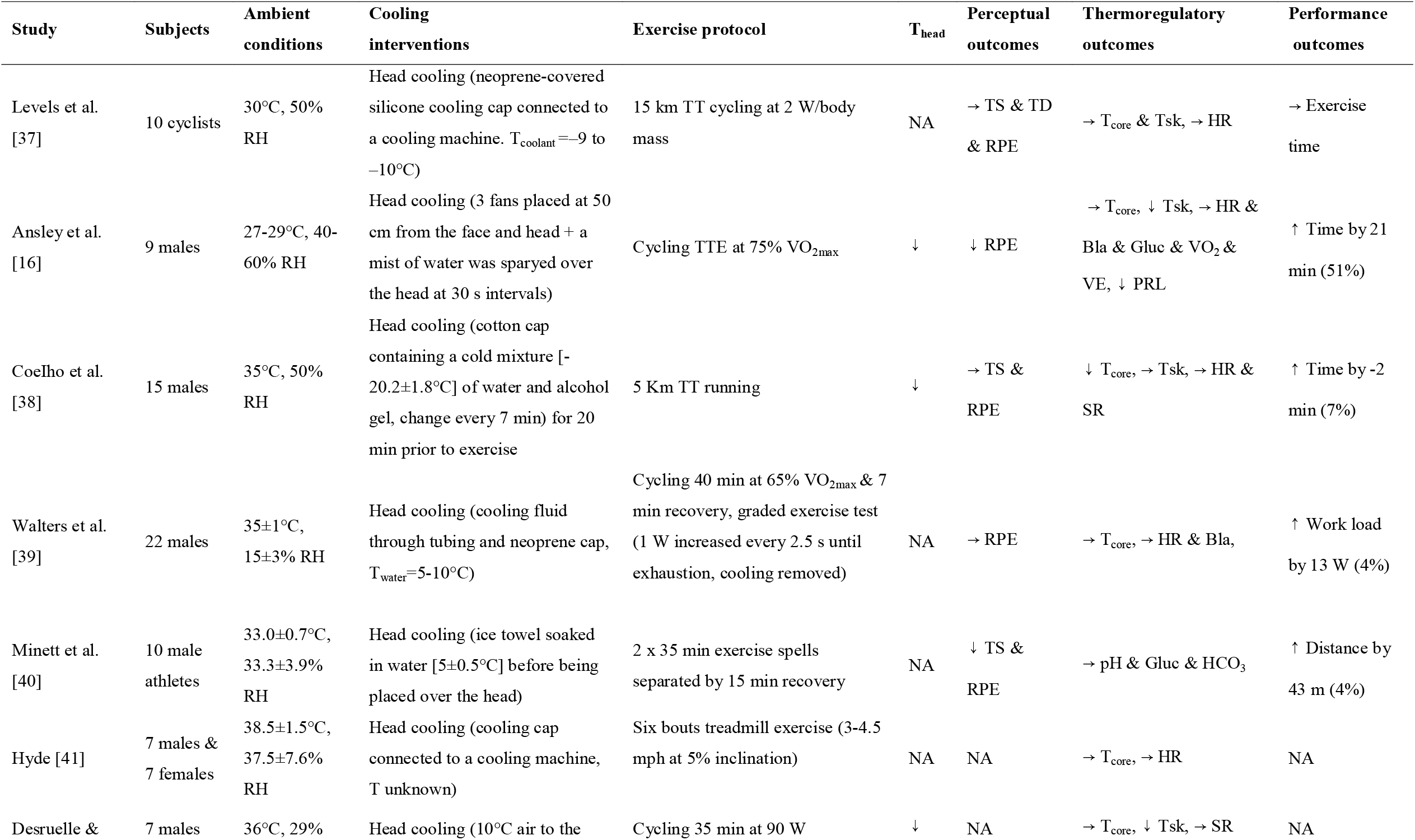
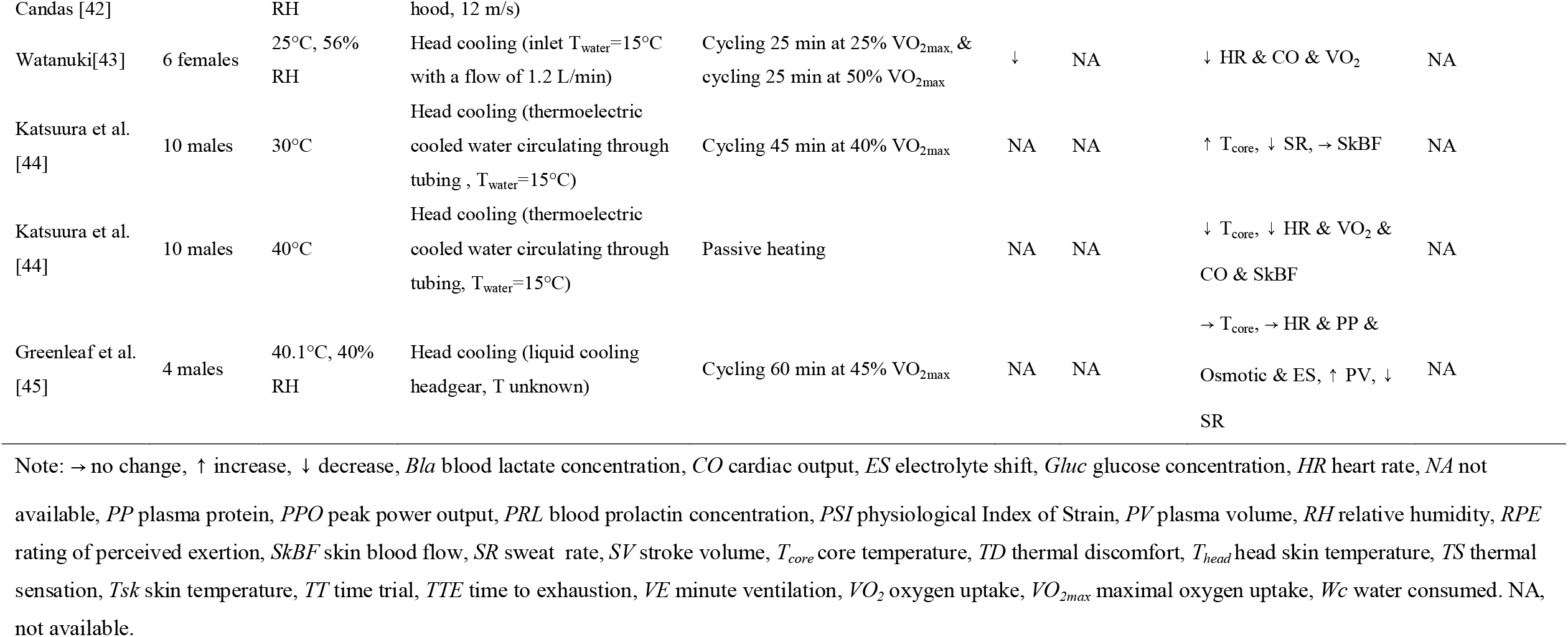
Effect of head cooling on thermoregulatory responses and physical performance in the heat.

| Study | Subjects | Ambient conditions | Cooling interventions | Exercise protocol | T<sub>head</sub> | Perceptual outcomes | Thermoregulatory outcomes | Performance outcomes |
| --- | --- | --- | --- | --- | --- | --- | --- | --- |
| Levels et al. [37] | 10 cyclists | 30°C, 50% RH | Head cooling (neoprene-covered silicone cooling cap connected to a cooling machine. T<sub>coolant</sub> = -9 to -10°C) | 15 km TT cycling at 2 W/body mass | NA | → TS & TD & RPE | → T<sub>core</sub> & Tsk, → HR | → Exercise time |
| Ansley et al. [16] | 9 males | 27-29°C, 40-60% RH | Head cooling (3 fans placed at 50 cm from the face and head + a mist of water was sprayed over the head at 30 s intervals) | Cycling TTE at 75% VO<sub>2max</sub> | ↓ | ↓ RPE | → T<sub>core</sub>, ↓ Tsk, → HR & Bla & Gluc & VO<sub>2</sub> & VE, ↓ PRL | ↑ Time by 21 min (51%) |
| Coelho et al. [38] | 15 males | 35°C, 50% RH | Head cooling (cotton cap containing a cold mixture [-20.2±1.8°C] of water and alcohol gel, change every 7 min) for 20 min prior to exercise | 5 Km TT running | ↓ | → TS & RPE | ↓ T<sub>core</sub>, → Tsk, → HR & SR | ↑ Time by -2 min (7%) |
| Walters et al. [39] | 22 males | 35±1°C, 15±3% RH | Head cooling (cooling fluid through tubing and neoprene cap, T<sub>water</sub> = 5-10°C) | Cycling 40 min at 65% VO<sub>2max</sub> & 7 min recovery, graded exercise test (1 W increased every 2.5 s until exhaustion, cooling removed) | NA | → RPE | → T<sub>core</sub>, → HR & Bla, | ↑ Work load by 13 W (4%) |
| Minett et al. [40] | 10 male athletes | 33.0±0.7°C, 33.3±3.9% RH | Head cooling (ice towel soaked in water [5±0.5°C] before being placed over the head) | 2 x 35 min exercise spells separated by 15 min recovery | NA | ↓ TS & RPE | → pH & Gluc & HCO<sub>3</sub> | ↑ Distance by 43 m (4%) |
| Hyde [41] | 7 males & 7 females | 38.5±1.5°C, 37.5±7.6% RH | Head cooling (cooling cap connected to a cooling machine, T unknown) | Six bouts treadmill exercise (3-4.5 mph at 5% inclination) | NA | NA | → T<sub>core</sub>, → HR | NA |
| Desruelle & | 7 males | 36°C, 29% | Head cooling (10°C air to the | Cycling 35 min at 90 W | ↓ | NA | → T<sub>core</sub>, ↓ Tsk, → SR | NA |
| Candas [42] |  | RH | hood, 12 m/s) |  |  |  |  |  |
| Watanuki[43] | 6 females | 25°C, 56% RH | Head cooling (inlet T<sub>water</sub>=15°C with a flow of 1.2 L/min) | Cycling 25 min at 25% VO<sub>2max</sub>, & cycling 25 min at 50% VO<sub>2max</sub> | ↓ | NA | ↓ HR & CO & VO<sub>2</sub> | NA |
| Katsuura et al. [44] | 10 males | 30°C | Head cooling (thermoelectric cooled water circulating through tubing , T<sub>water</sub>=15°C) | Cycling 45 min at 40% VO<sub>2max</sub> | NA | NA | ↑ T<sub>core</sub>, ↓ SR, → SkBF | NA |
| Katsuura et al. [44] | 10 males | 40°C | Head cooling (thermoelectric cooled water circulating through tubing, T<sub>water</sub>=15°C) | Passive heating | NA | NA | ↓ T<sub>core</sub>, ↓ HR & VO<sub>2</sub> & CO & SkBF | NA |
| Greenleaf et al. [45] | 4 males | 40.1°C, 40% RH | Head cooling (liquid cooling headgear, T unknown) | Cycling 60 min at 45% VO<sub>2max</sub> | NA | NA | → T<sub>core</sub>, → HR & PP & Osmotic & ES, ↑ PV, ↓ SR | NA |

Note: → no change, ↑ increase, ↓ decrease, *Bla* blood lactate concentration, *CO* cardiac output, *ES* electrolyte shift, *Gluc* glucose concentration, *HR* heart rate, *NA* not available, *PP* plasma protein, *PPO* peak power output, *PRL* blood prolactin concentration, *PSI* physiological Index of Strain, *PV* plasma volume, *RH* relative humidity, *RPE* rating of perceived exertion, *SkBF* skin blood flow, *SR* sweat rate, *SV* stroke volume, $T_{\text{core}}$ core temperature, *TD* thermal discomfort, $T_{\text{head}}$ head skin temperature, *TS* thermal sensation, *Tsk* skin temperature, *TT* time trial, *TTE* time to exhaustion, *VE* minute ventilation, $\text{VO}_2$ oxygen uptake, $\text{VO}_{2\text{max}}$ maximal oxygen uptake, *Wc* water consumed. NA, not available.

## 5 Facial cooling and exercise performance

The facial cooling and its performance outcomes are shown in Table 3. Of all 9 included studies, 8 of them examined either the forehead or the facial temperature. Five studied [12, 46–49–51] also examined the local perceptual sensations (thermal sensation and thermal discomfort). Similar to the effect of neck and head cooling on the neck and head regions, facial cooling could greatly enhance the thermal sensation at the facial area and thermal discomfort at the face could be greatly alleviated as well. For physiological outcomes, although two studies [12, 46] have suggested that facial cooling seemingly enhance performance in the heat (Table 3), facial cooling may not fully render ergogenic benefits in the heat for both endurance and team sport athletes in the heat. This is given by the fact that there are no studies available in team sport athletes and facial cooling only reduced RPE at the end of the stage [50]. Such small decrement of RPE may not be large enough to elicit behavioral adaptation in highly trained subjects as their perceptual response is different from healthy population [61]. In particular, Stevens et al. [46] indicated that even in moderately trained subjects, facial cooling only improved running speed at the first 2-km but it was soon nullified thereafter. Furthermore, no studies are available to show whether facial cooling can potentially extend the time to exhaustion in well trained endurance athletes or with different levels of aerobic fitness. Those evidence from above support the notion that facial cooling may not be applied to well-trained subjects.

**Table 3.**
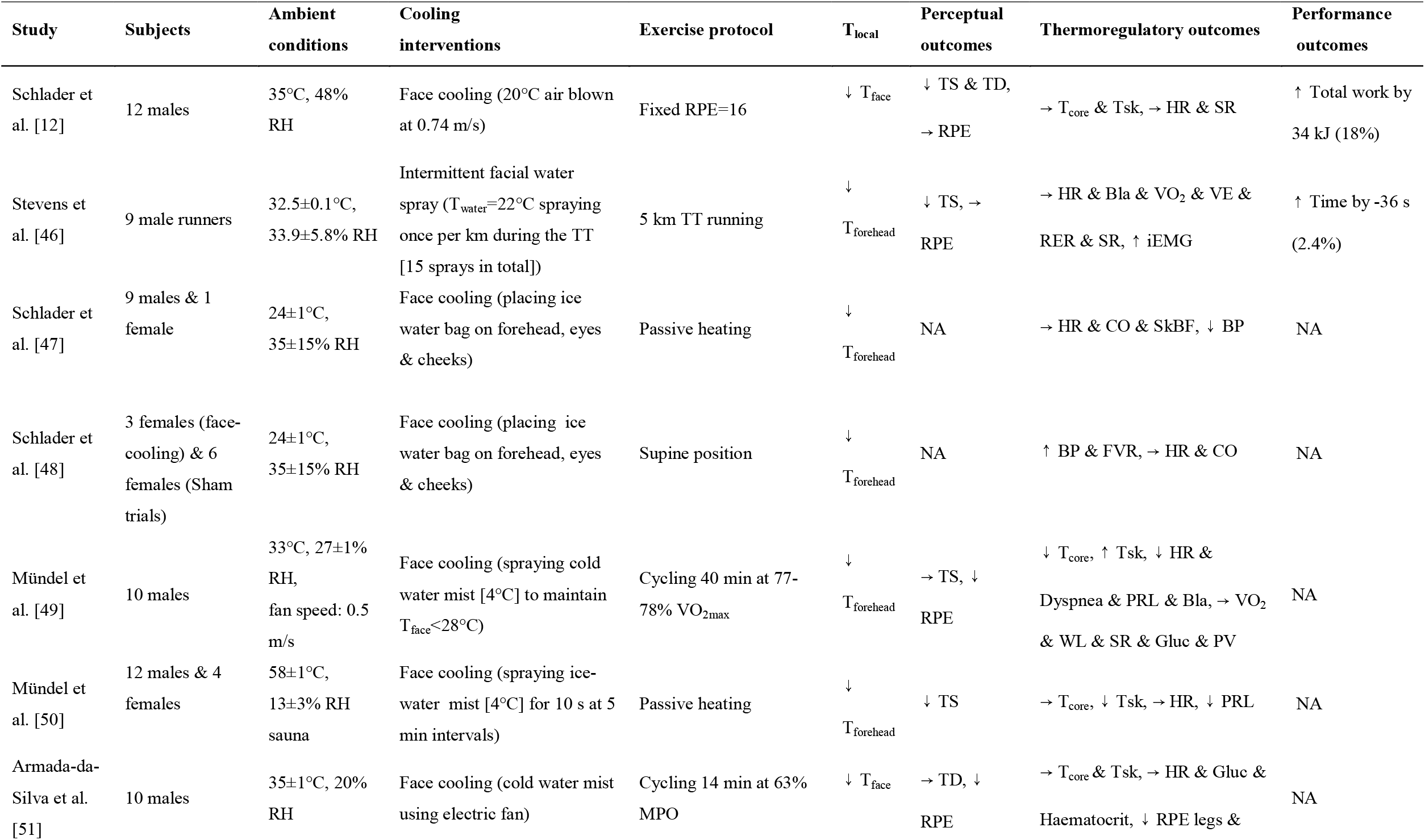
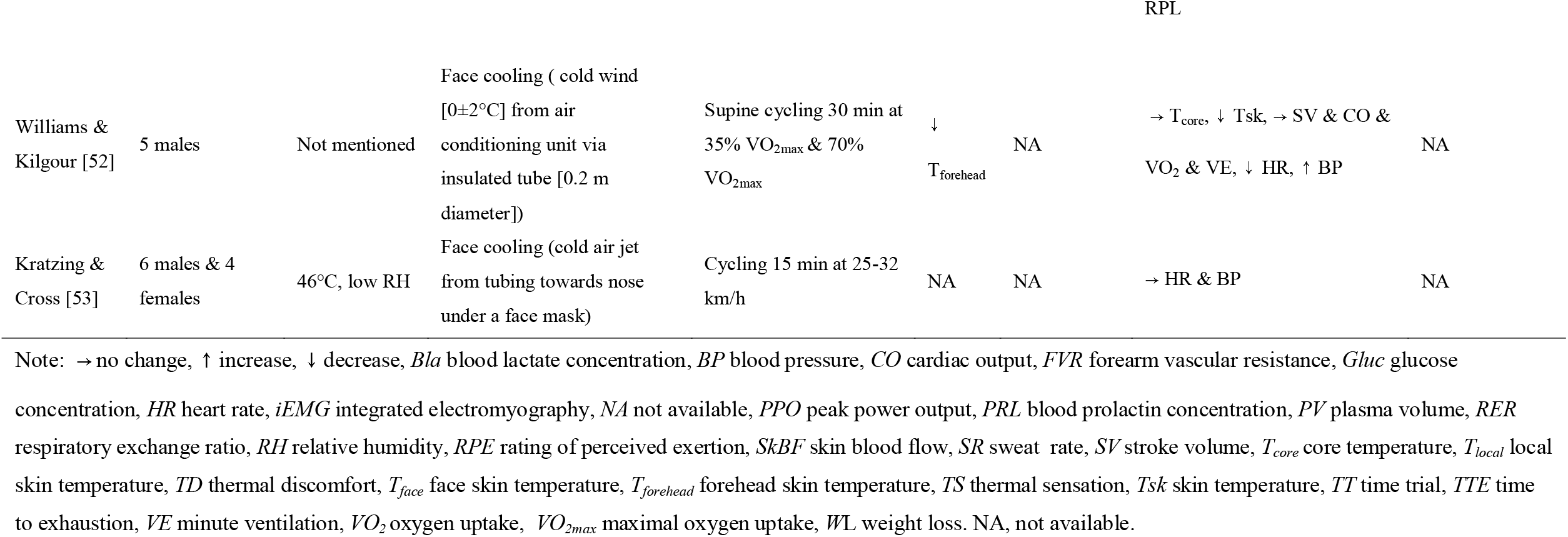
Effect of facial cooling on thermoregulatory responses and physical performance in the heat.

From the practical point of view, facial cooling also has a poor real-world implication to both endurance and team sport athletes as it is difficult to employ this during exercise. It is well noted that per cooling is just as important as pre-cooling especially when the environment is hot and humid. Therefore, facial cooling is not an ideal cooling method during exercise in a hot humid environment.

## 6 Combined head, neck and/or facial cooling & exercise performance

The impact of combined facial/head and neck cooling on the sports and exercise performance is presented in Table 4 [54–60]. Obviously, the combined hear/face and neck cooling could largely improve the thermal sensation and thermal discomfort at the cooling regions [54–59]. The effect of combined effect of either head and neck or facial and head cooling on exercise performance in the heat remains unclear and may not actually render any ergogenic benefits compared to neck cooling along despite a larger total cooling area in the combined cooling strategy as compared to the single cooling approach being applied to the face, neck or the head [61]. This is given by the fact that current literatures have yet to address the combined effect of either head and neck cooling or facial and head cooling on exercise performance as compared neck cooling along in the heat for both endurance and team sport athletes. Furthermore, by summarizing the current literatures from above (Table 1, Table 2 and Table 3), it is suggested that the combined cooling may not actually render any ergogenic benefits as the effect of both head and facial cooling on both endurance and repeated sprint performance remains equivocal. Lastly, from the practical perspective, combined cooling is considered having less real-world application than neck cooling along as it is relatively difficult to implement this cooling intervention during exercise in the heat.

**Table 4.**
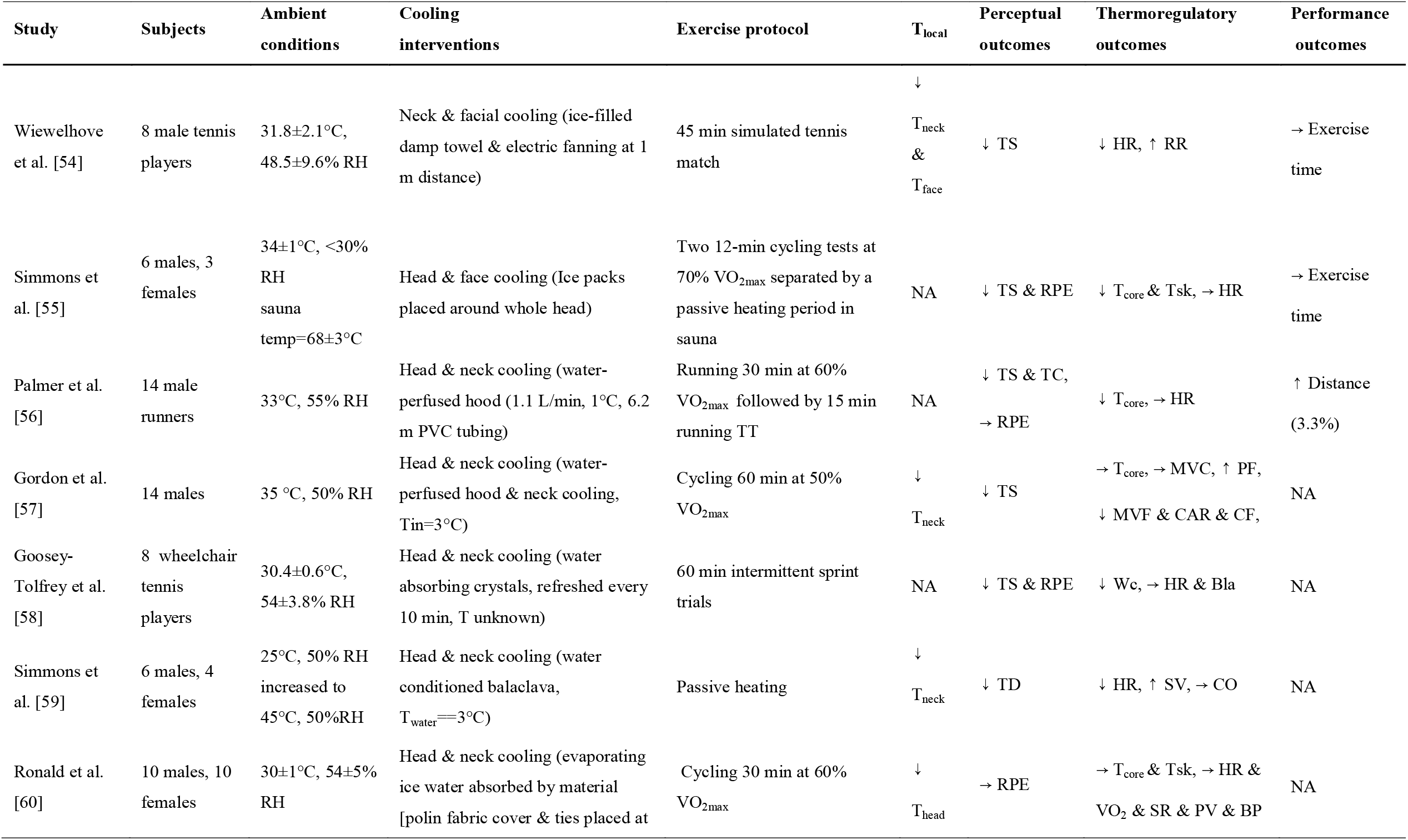
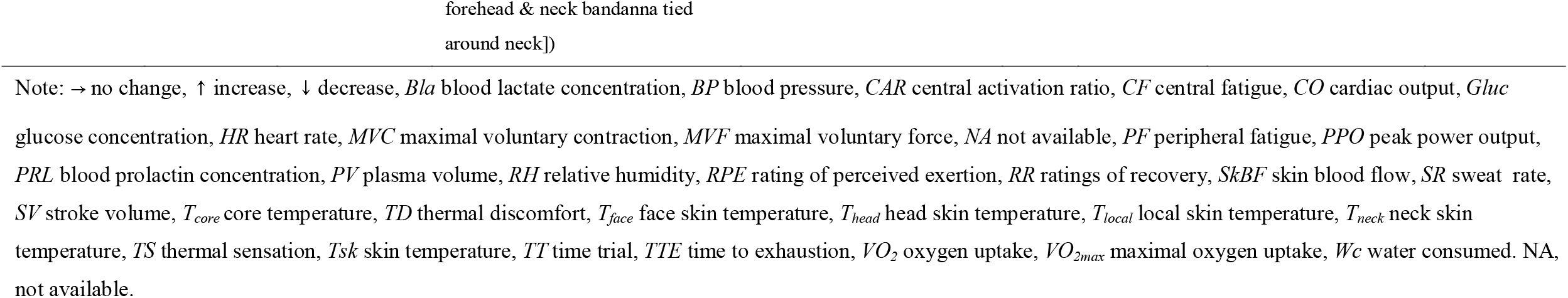
Effect of combined neck, head and facial cooling on thermoregulatory responses and physical performance in the heat.

Although the combined cooling intervention may not be suitable as a per cooling strategy for both endurance and team sport athletes in the heat, this cooling intervention may be effective as a post-cooling strategy after performing exercise in the heat as this may be able to compensate the withdrawal of thermoeffector and thereby reducing the magnitude of post-exercise hyperthermia. In particular, occupational workers with protective clothing or American football players with multiple body armor may be beneficial from this intensive cooling intervention as compared to neck cooling along. However, this separate and combined effect have not been investigated and therefore warrants further investigation.

## 7 Practical considerations

By summarizing the current literatures to date (Tables 1,2, 3 & 4), neck cooling is preferred over head or facial cooling as it can be used before and during exercise in the heat for both endurance and team sport athletes [62]. Besides, head cooling is preferred over facial cooling as it can also be used as a pre and per cooling strategy in the heat. It is worth mentioning that the selection of cooling temperature from the neck cooling collar or cooling helmet needs to be individualised as each individual will have their own preferred cooling temperature but the cooling temperature needs to be great enough to alter the RPE whilst without causing further rise of core temperature. Furthermore, it is suggested that the endurance athletes should also utilise either the neck or head cooling intervention upon the completion of their race as it is necessary to compensate the effect of post-exercise hyperthermia especially when the heat stress environment is hot and humid [63].

Lastly, it is suggested that the cooling materials inside the cooling collar or the helmet should consider phase change materials (including ice) rather than the gel refrigerant because phase change materials provide stable cooling intensity than the gel refrigerant during phase change. Though soft gel could provide good flexibility when being applied to the body surface, its temperature changes throughout the entire application period. Particularly, soft gel refrigerant provides the most powerful cooling intensity at the very beginning but this cooling power gradually reduces with the time. In addition, the mass of phase change materials should be large in order to provide athletes an extended cooling duration during exercise. Other cooling strategies such as the use of wearable cooling fans and a liquid cooling neck collar may be applicable, their actual performance on the improvement of performance in exercising athletes requires further investigation, however.

## 8 Directions for Future Research

Whilst current studies have eloquently addressed the effect of neck, head, facial as well as the combined head & neck cooling on both endurance and team sport performance, there are three major unexplored issues that warrants further investigation. First, current studies did not specifically incorporate the effect of different cooling modes on local and whole-body thermal discomfort, thermal pleasant and skin wettedness in their research design. It is well known that those thermal perceptions could influence RPE and thereby affecting our physical performance in the heat [12, 64]. Furthermore, those perceptual variables are important to dictate our thermoregulatory behavior [12, 65, 66] and therefore it would be novel to investigate whether cooling per se would mask the signal to initiate our thermoregulatory behavior, which has the unlimited capacity to regulate our body temperature.

Second, previous studies do not consider the role of aerobic fitness on the effect of neck, head, facial and combined head & neck cooling during the self-paced exercise in the heat. It is well known that well trained populations are more accustom to being thermally discomfort [67] than untrained individual when exercising in the heat and therefore additional cooling may not be ergogenic as compared to untrained individual. Indeed, previous studies mainly target endurance or team sport athletes and this could be the reason why some studies did not find any performance enhancement by using neck cooling during exercise.

Lastly, rationale on the selection of cooling devices in a given sports and exercise setting has not yet been well investigated. For instance, the cooling temperature of the device varies considerably ranging from 21 °C to −80 °C and such variation of cooling temperature would undoubtfully alter our thermal perceptions and hence our exercise performance in the heat. Furthermore, to date, the selection of those cooling temperatures is not evidenced based, instead it is chosen without any supporting evidence from either the thermoregulatory or the perceptual response in the heat. Also, the mass of cooling materials, the type of cooling strategy (e.g., air cooling, phase change material cooling, evaporative cooling, & liquid cooling), the design of the cooling devices (e.g., packing material, insulation between the cooling material and human skin, & contact area) are far from clear. Those factors could greatly affect the cooling intensity as well as the cooling duration of the selected device. Thereby, their actual impact on the performance enhancement while exercising in various heat conditions could vary greatly. Therefore, those aforementioned issues warrant further investigation as this would extend our understanding on the effect of per cooling in both compensable or in uncompensable heat stress environment.

## 9 Conclusions

Neck, facial and head cooling could significantly reduce local skin temperature and thereby improves local perceptual sensations including thermal sensation and thermal discomfort on athletes while exercising in the heat. Neck cooling is an ideal per-cooling strategy than head or facial cooling for both endurance and team sport athletes. Furthermore, head cooling is preferred than facial cooling for endurance athletes but its efficacy on team sport athletes remains to be explored. Furthermore, it remains unclear whether the combined head, neck and/or facial cooling could bring a synergy effect on sports and exercise performance in the heat. Future investigations should extensively examine the potential of neck, head and facial cooling to enhance performance of male and female athletes while performing various types of sport and exercise activities in the heat. Besides, the design of powerful but portable head, neck and facial cooling systems is urgently required due to the great need to dissipate metabolic heat production during intensive sports activities.

## Data Availability

Data are available on request from the authors.

## Author contributions

Original idea: FW; Development and formulation of concept: FW, THL, YC & BY; Draft: YC, THL, FW; Critical revision: THL, FW & BY.

